# Comorbidity of self-harm and disordered eating in young people: Evidence from a UK population-based cohort

**DOI:** 10.1101/2020.07.08.20148908

**Authors:** Naomi Warne, Jon Heron, Becky Mars, Paul Moran, Anne Stewart, Marcus Munafò, Lucy Biddle, Andy Skinner, David Gunnell, Helen Bould

## Abstract

Self-harm and eating disorders are often comorbid in clinical samples but their co-occurrence in the general population is unclear. We assessed the co-occurrence of self-harm and disordered eating behaviours at age 16 and 24 years in 3384 females and 2326 males from a UK population-based cohort. There was substantial overlap at both time points: almost two-thirds of 16-year-old females, and two-in-five 24-year old males who self-harmed also reported disordered eating. Among young people, self-harm may signal the occurrence of disordered eating and vice versa. It is important to screen for both sets of difficulties to provide appropriate treatment.

## Introduction

Eating disorders and self-harm are serious health problems in young people, associated with significant functional impairment and mortality(1,2). They are phenotypically distinct - eating disorders involve weight-control behaviours, abnormal eating and over-evaluation of weight and shape(2), whereas self-harm involves intentionally harming oneself, with or without suicidal intent(1). However, they commonly co-occur in clinical populations: 14-68% of patients with eating disorders report self-harm and 54-61% of self-harm patients report eating disorders(3). This comorbidity may reflect common risk factors, such as emotion dysregulation or impulsivity(3). Few studies assess comorbidity of disordered eating and self-harm in non-clinical samples. Such research is important as only a small proportion of individuals who self-harm or have disordered eating present to clinical services(1,2).

In non-clinical samples, 50.8% of female university students who self-harmed reported a possible eating disorder, and 20.1% with a possible eating disorder reported self-harm(4). A community sample of 14-to 19-year-olds(5) found higher levels of disordered eating in those who had self-harmed in the last year, compared to those who had not. Although these studies are informative, unselected samples are required to understand how self-harm and disordered eating co-occur in the general population over the adolescent to young adult age range, during which rates of both may vary(1,2,6).

In this study, we examine the co-occurrence of self-harm and disordered eating at 16 and 24 years in a UK population-based cohort.

## Methods

Data were from a prospective birth cohort: The Avon Longitudinal Study of Parents and Children (ALSPAC)(7,8). ALSPAC recruited pregnant women with expected delivery dates between April 1991 and December 1992 in Avon, UK (core sample n=13,988 alive at 1 year). Ethics approval for the study was obtained from the ALSPAC Ethics and Law Committee and the Local Research Ethics Committees. The study website contains details of ALSPAC data: http://www.bristol.ac.uk/alspac/researchers/our-data.

We conducted our analysis on an imputed dataset of 5710 individuals (3384 females, 2326 males) who completed self-harm and disordered eating questionnaires at age 16 and/or 24 years (see Supplementary Material). *Disordered eating behaviours* in the last year were assessed on adapted Youth Risk Behaviour Surveillance System(9) questions. Behaviours included *fasting* (not eating for at least one day), *purging* (vomiting or taking laxatives/other medicines), and *excessive exercise* (that frequently interfered with daily routine/work) in order to lose weight or avoid gaining weight, as well as *binge-eating* (eating a very large amount of food, with loss of control, in a short period of time). Behaviours were considered present if endorsed at any frequency in the last year. Our primary variable of interest was *any disordered eating* (any of the aforementioned behaviours); we also present data for individual behaviours and *any disordered eating at DSM-5 frequency* (any of the behaviours at least once a week). For *self-harm* in the last year, participants were asked adapted Child and Adolescent Self-Harm in Europe study(10) questions on whether they had harmed themselves on purpose in any way (regardless of suicidal intent), and when this occurred. Questions and variable coding are available in Table S1.

Analyses were a priori stratified by gender. At each age, we assessed 1) the proportion of individuals reporting disordered eating and self-harm in the sample; 2) what proportion of individuals with disordered eating also reported self-harm (compared to those without disordered eating); and 3) what proportion of individuals who self-harmed also reported disordered eating (compared to those not self-harming). Results were consistent between imputed and complete case analysis (Tables S4 and S5).

## Results

At age 16, 32.7% of females and 7.6% of males reported disordered eating in the past year, whereas 15.3% of females and 5.4% of males reported past-year self-harm (Table S2).

Prevalence of past-year disordered eating increased for both females (36.9%) and males (19.2%) at age 24 but self-harm decreased in females (9.8%) and males (5.1%).

Co-occurrence of self-harm and disordered eating was common (Figure 1). In those reporting any disordered eating at age 16, 29.9% of females and 23.7% of males also self-harmed in the last year, compared to 8.3% of females and 4.0% of males without disordered eating. At age 24, 16.1% of females and 11.1% of males with any disordered eating also self-harmed, compared to 6.0% of females and 3.6% of males without disordered eating. The eating behaviours associated with the highest levels of self-harm in females were purging at 16 (45.4%) and excessive exercise at 24 (23.2%); in males it was purging at 16 (34.4%) and fasting at 24 (17.9%).

**Figure 1.**
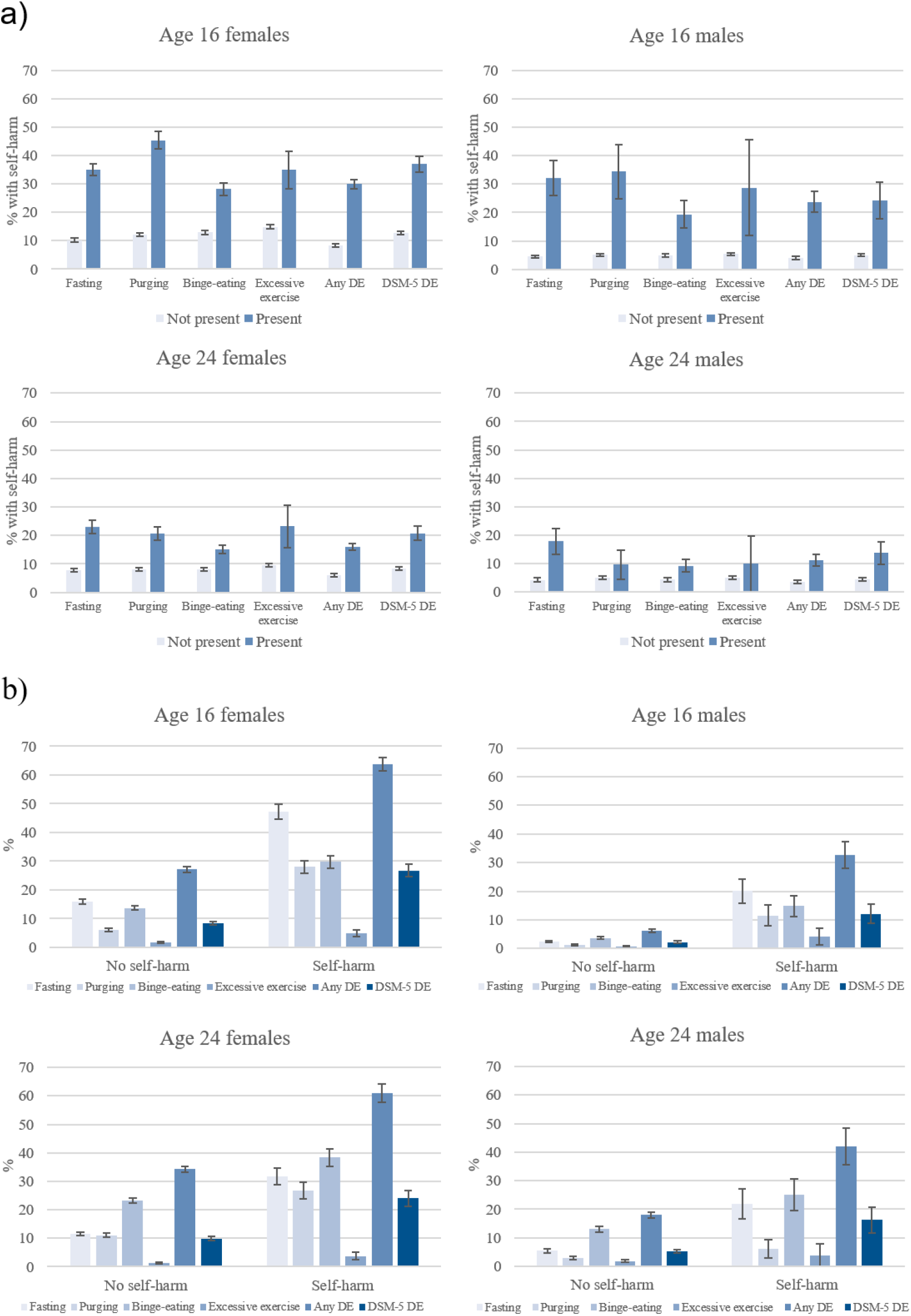
Co-occurrence of self-harm and disordered eating behaviours. Panel a) Self-harm in individuals with or without disordered eating behaviours. Panel b) Disordered eating behaviours in individuals with or without self-harm. Any DE = any disordered eating (fasting, purging, binge-eating, or excessive exercise); DSM-5 DE = disordered eating (fasting, purging, binge-eating, or excessive exercise) at least once a week (DSM-5 frequency). Error bars indicate standard error.

At age 16, 63.7% of females and 32.7% of males who self-harmed also reported disordered eating, compared to 27.1% of females and 6.1% of males who did not self-harm. At 24, 60.9% of females who had self-harmed (versus 34.3% who had not) and 41.9% of males who had self-harmed (versus 18.0% who had not) reported disordered eating. In those with self-harm, the highest proportions of concurrent disordered eating behaviours were fasting (47.2%) for 16-year-old females, binge-eating (38.4%) for 24-year-old females, fasting (20.1%) for 16-year-old males, and binge-eating (25.1%) for 24-year-old males.

## Discussion

In a UK population-based cohort, we found that self-harm and disordered eating behaviours were common, and commonly co-occurred in young people. The prevalence of disordered eating was notably high among females who reported self-harm, affecting nearly two-thirds of young women at each time point. Furthermore, two-in-five 24-year-old males who self-harmed also reported disordered eating.

This high rate of co-occurrence is consistent with findings from selected clinical(3), university student(4), and adolescent(5) samples. Our study extends findings to young adults in the community. This is particularly important since the majority of young people with self-harm or disordered eating do not seek help(1,2). Consistent with the previous literature(3–5), we found higher rates of disordered eating among those reporting self-harm than vice versa.

This comorbidity may be due to shared risk factors that contribute to the development of both disordered eating and self-harm. A number of potential risk factors (such as impulsivity, emotion dysregulation and dissociation(3)) have been suggested based on evidence from clinical samples. However, given the small proportions of those with self-harm and disordered eating who present to clinics(1,2), longitudinal studies in population-based samples are needed to assess factors that precede self-harm and disordered eating behaviours. Such research could facilitate early identification of those at high risk of developing self-harm and/or disordered eating and identify modifiable targets for prevention and intervention measures.

The strengths of the current study include the large population-based sample and examination of multiple types of disordered eating over this important life period. There are limitations: firstly, the small number of males reporting behaviours and lack of data at other ages means results may not generalise. Secondly, questions identifying disordered eating may have excluded those with milder, although significant, symptoms. Thirdly, some individuals may self-define their disordered eating as self-harm; we were not able to differentiate when this was the case. Finally, we used an imputed dataset under the assumption data are missing at random, which if not true, could mean results are biased.

In summary, we found substantial comorbidity between self-harm and disordered eating in young adults in the general population. Health professionals should be aware of this comorbidity and ensure that young people presenting with either self-harm or disordered eating are asked about both behaviours in order to provide appropriate treatment/management.

## Data Availability

ALSPAC data is available to researchers through an online proposal system. Information regarding access can be found on the ALSPAC website: http://www.bristol.ac.uk/media-library/sites/alspac/documents/researchers/data-access/ALSPAC_Access_Policy.pdf

## Funding

This work was supported by funding from the Medical Research Council/Medical Research Foundation (MRC/MRF grant number MR/S020292/1).

Becky Mars, David Gunnell and Paul Moran are part-funded by the NIHR Biomedical Research Centre at University Hospitals Bristol NHS Foundation Trust and the University of Bristol. Paul Moran and Lucy Biddle are part-funded by NIHR Applied Research Collaboration (ARC) West. Andy Skinner is funded by a UKRI Innovation Fellowship from Health Data Research UK to Andy Skinner (MR/S003894/1). The views expressed in this correspondence are those of the authors and not necessarily those of the National Health Service, the National Institute for Health Research, or the Department of Health and Social Care.

The UK Medical Research Council and Wellcome (Grant ref: 217065/Z/19/Z) and the University of Bristol provide core support for ALSPAC. This publication is the work of the authors and Naomi Warne and Helen Bould will serve as guarantors for the contents of this paper. A comprehensive list of grants funding is available on the ALSPAC website (http://www.bristol.ac.uk/alspac/external/documents/grant-acknowledgements.pdf); This research was specifically funded by Wellcome Trust and MRC (Grant ref: 092731), NIH (Grant refs: MH087786-01 & R21MH109917), Wellcome Trust (Grant ref: GR067797MA) and NIHR (Grant ref: 1215-20011).

Acknowledgements

We are extremely grateful to all the families who took part in this study, the midwives for their help in recruiting them, and the whole ALSPAC team, which includes interviewers, computer and laboratory technicians, clerical workers, research scientists, volunteers, managers, receptionists and nurses.

## Author Contribution

HB, JH, BM, PM, ASt, MM, LB, ASk, and DG were involved in obtaining research funding for the project. All authors were involved in formulating the research question. NW and JH performed data analysis. All authors contributed to interpretation of results and writing of the final manuscript.

## Data availability

ALSPAC data access is through a system of managed open access. The steps below highlight how to apply for access to the data included in this paper and all other ALSPAC data.

1. Please read the ALSPAC access policy (PDF, 843kB) which describes the process of accessing the data and samples in detail, and outlines the costs associated with doing so.
2. You may also find it useful to browse our fully searchable [research proposals database](https://proposals.epi.bristol.ac.uk/), which lists all research projects that have been approved since April 2011.
3. Please [submit your research proposal](https://proposals.epi.bristol.ac.uk/) for consideration by the ALSPAC Executive Committee. You will receive a response within 10 working days to advise you whether your proposal has been approved.

If you have any questions about accessing data or samples, please (data) or (samples).

